# Family-centered postnatal training and adherence to newborn care practices in district hospitals in two Indian states: A quasi-experimental intervention study

**DOI:** 10.1101/2021.03.15.21252605

**Authors:** Sehj Kashyap, Amanda F. Spielman, Nikhil Ramnarayan, SD Sahana, Rashmi Pant, Baljit Kaur, N Rajkumar, Ramaswamy Premkumar, Shahed Alam, Seema Murthy

## Abstract

**Background and Objectives:** Globally, 2.5 million newborns die within the first month of life annually. The majority of deaths occur in low- and middle-income countries (LMICs), and many of these deaths happen at home. The study assessed if the Care Companion Program (CCP) an in-hospital, skills-based training given to families improves post-discharge maternal and neonatal health outcomes.

**Methods:** This quasi-experimental pre-post intervention study design compared self-reported behavior and health outcomes among families before and after the CCP intervention. Intention to treat analysis included families regardless of their exposure to the intervention. Mixed effects logistic regression model, adjusted for confounders, was fit for all observations. Effects were expressed as Relative Risks (RR) with 95% Confidence Intervals (CI).

**Results:** At 2-weeks post-delivery, telephone surveys were conducted in the pre (n = 3510) and post-intervention (n = 1474) groups from 11 district hospitals in the states of Karnataka and Punjab. The practice of dry cord care improved significantly by 4%, (RR = 1.04, 95%CI [1.04,1.06]) and skin to skin care by 78% (RR=1.78, 95%CI [1.37,2.27]) in the post-intervention group as compared to pre-intervention group. Furthermore, newborn complications reduced by 16% (RR=0.84, 95%CI [0.76,0.91]), mother complications by 12% (RR=0.88, 95%CI [0.79,0.97]) and newborn readmissions by 56% (RR=0.44, 95%CI [0.31,0.61]). Outpatient visits increased by 27% (RR=1.27, 95%CI [1.10,1.46]). However, outcomes of breastfeeding, mother’s diet, hand-hygiene, and process indicator of being instructed on warning signs were not different.

**Conclusion:** Postnatal care should incorporate pre-discharge multi-pronged training of families to improve essential maternal and newborn care practices. The CCP model runs on a public-private partnership and is integrated into existing health systems. Our findings demonstrate that it is possible to improve outcomes through a family-centered approach in India. The CCP model can be integrated into formalised hospital processes to relieve overburdened healthcare systems in LMIC settings.

**ARTICLE SUMMARY:** *Strengths and Limitations of this study:*

1. Study in 11 public health settings in 2 large Indian States and having large sample size
2. Multiple health behaviours measured post-discharge during the neonatal period
3. Quasi-experimental study design allows for historical control group in the same setting
4. Primary limitation is that all behaviours are self-reported
5. Random allocation was not feasible for this group intervention

## INTRODUCTION

Reducing neonatal morbidity and mortality remains a key goal of health systems in low- and middle-income countries (LMICs) where 2.5 million neonatal deaths occur annually[1]. India bears 27% of the global burden of neonatal deaths; 750,000 newborns die in India within the first month of life annually[2].

Although institutional deliveries have nearly doubled in India over the past decade[3], inadequate follow-up has created several gaps in care. Families often miss coming back for scheduled follow-ups, with about 40% dropping out by the first follow-up visit in LMICs[4]. Additionally, many patients fall through the safety net as health workers are not able to reach and provide care for the newborns at home[5]. Thus irrespective of the place of delivery, most neonatal deaths occur in the home setting[6]. For many families, facility-based childbirth is the only window of opportunity to equip families with the knowledge and skills needed to care for their newborns.

Postnatal education, particularly predischarge education has been identified as a “low-hanging fruit” to improve newborn outcomes and reduce mortality by improving the adoption of newborn care practices[7,8]. But key limitations have been identified in the implementation of such programs. The majority of programs focus on singular health topics, primarily breastfeeding, or impart education to only one family member, most often the mother. These limitations need to be overcome to address unmet educational needs[8].

Hospitals in LMICs can play a key role in imparting postnatal education[4], but without established procedures the delivery of critical health education in-hospital is limited or missed altogether. A recent survey in district hospitals in India found less than half the mothers reported the receipt of any amount of education post-delivery and before discharge[8]. Moreover, low health literacy, socio-cultural diversity, power dynamics, and language barriers affect patient provider relationships, thereby adding to the difficulty in understanding the advice given by doctors and nurses. Often, families are not prepared for their roles as primary caregivers and essential newborn care practices that could save lives and avert suffering are not followed at home[9].

### Program Description

Recently, eleven district hospitals in two states in India adopted a novel approach to patient education with families in hospitals being trained on essential newborn care, post-delivery. Noora Health inspired Care Companion Program (CCP) was implemented as a public-private partnership undertaking in two states of India. The program has an evidence-based curriculum for postnatal counselling, covering multiple behaviors for improving neonatal and maternal health[10] Skills to facilitate behavioral change and promote healthy outcomes are taught; they include family’s understanding and practice of key healthy behaviors, healthcare system’s support for family engagement, health care seeking by the families, and complications. Mothers and families are taught these skills in group sessions held in the hospitals.

Tools and methods to teach this curriculum were created using a human-centered design process including needs-finding interviews with families and discussions with local and regional health experts. The CCP trained nurses and counselors in the district hospitals primarily in health communication skills to engage their audiences[10]. Trainers were taught adult learning principles and methods of empathic medical communication. Train-the-trainer sessions took place over two 8-hour days. Visuals and materials were tailored to the cultural practices of local populations using field testing, key informant interviews and iterative design. Final sign-off on the materials was granted by the Departments of Health in the respective states as well as District Surgeons and Medical Officers of each hospital. The materials included videos which are played on a television monitor installed in the postnatal wards, flipcharts, dolls for role-play, and hand-outs. A rotational district specific and ward specific roster was created in order to cover all deliveries and these trainers take turns in taking group classes for families.

The training session involved formal group sessions with families in postnatal wards. The frequency of these classes depended on patient turn-over-rates in the particular ward of the facility. For instance, the classes in postnatal wards of vaginal delivery mothers were more frequent (twice a week or more) than in cesarean section wards (once a week or more). On average, each facility ran 1-2 sessions per week. The primary SMART aim of the CCP is to improve patients’ post-discharge outcomes, reduce complications and increase families’ adherence to recommended newborn care practices. Our objective in the study was to assess the effect of CCP on family’s reported adoption of newborn care practices and newborn outcomes in the neonatal period.

## METHODS

### Study Design

We conducted a quasi-experimental study where a pre-intervention group received standard of care (SoC) and served as an historical control and the post-intervention group receiving the CCP served as the intervention arm. All outcomes were self-reported by the mother or a primary caregiver who lived with the mother and were collected at 2-weeks post-delivery via telephone survey. We were interested in the overall effect of the program at the systems level. Hence intention to treat analysis was applied so that families were included in the analysis regardless of their individual level of exposure to postnatal education.

Eleven district hospitals were selected by the governments of Punjab (5 hospitals) and Karnataka (6 hospitals) to launch the Care Companion Program as a pilot program in the state. These hospitals have delivery rates ranging from 100-800 deliveries per month and are the primary government hospitals in their respective districts.

Pilot district hospitals were selected based on need for the program as determined by the state governments and willingness among the district hospital leads to implement the program. The CCP was launched in July 2017 and by August 2017 all hospitals were running the CCP. Families in the pre intervention group were recruited between May and June 2017. And for the post intervention group, between August and October 2017.

### Sample

Field investigators created a list of deliveries each day referencing the hospital’s delivery registers. Using survey software, we selected a random sample of women who delivered a newborn in the hospital during the study timeframe. Field investigators checked for inclusion and exclusion criterion by talking to the families and using individual patients’ chart data and developed a final list. The study was powered to detect a 19% improvement in exclusive breast feeding (EBF) practices at 6-weeks. Assuming 30% EBF at 6-weeks for the SoC group, to detect 19% improvement in EBF at 6-weeks for the CCP group, a sample size of at least 1425 participants in each group would be sufficient at 5% level with 90% power.

### Inclusion and exclusion criteria

All women who delivered a live newborn in the selected district hospitals during the study recruitment were eligible and included in the study sampling frame. Families were excluded from participating if baby or mother died during hospital stay or by the time of the survey call, mother was younger than 18 years, no one living near the mother had access to a telephone, no one living with the mother spoke one of the multiple languages the surveyors spoke, mother or baby were transferred to another hospital during their stay in the hospital, medicolegal cases and if the mother left the hospital before data collectors could collect telephone numbers.

### Ethics and consent

Participants provided oral informed consent in their primary language for participation. All surveys were recorded. The study was approved after review by local health authorities. ACE Independent Ethics Committee, (DCGI Reg. No. ECR/141/Indt/KA/2013) provided approval for the study.

### Patient and public involvement

Study questionnaires were piloted with the target population prior to the start of study data collection and based on the pilot feedback. The instrument was fine-tuned for the comfort and understanding of the patient population.

Once published, participants will be informed of the results through a dedicated section in the website (www.noorahealth.org), each hospital involved will be sent the results and the Government stakeholders at the state level will be informed.

### Measurements

Population based surveys like the Demographic and Health Survey [11] and National Family Health Survey [12] collect limited data about newborn care practices and neonatal and post-partum maternal health, only measuring breastfeeding practice and the occurrence of excessive vaginal bleeding or fever in the postpartum period. The study team developed and tested a phone-survey to additionally measure other newborn care practices that are shown to reduce newborn illness and death: skin to skin care, exclusive breastfeeding, infection prevention including handwashing and clean umbilical cord care, and care seeking for newborn illness. In addition, we chose to measure receipt of post-delivery instructions and problems like hospitalization and readmissions.

A phone-survey was developed in English and translated into Kannada, Hindi and Punjabi by certified translators. Given all measures were self-reported, several techniques were employed in the design of the survey to minimize desirability bias and recall bias: open-ended questions, small recall periods and specific question ordering. Where there was overlap in measurement, questions about EBF and postpartum complications were adapted from the NFHS and DHS surveys. The translated surveys were evaluated for face and content validity by survey managers fluent in the language and familiar with the study aims. Subsequently, the survey was tested with 20 families for understandability. Lastly, the survey was backtranslated by the study team that was bilingual and assessed.

The survey was electronically programmed into SurveyCTO, a tool for digital data collection. Required responses, response validity checks and skip patterns were used to ensure completeness of data; thus, only missing data occurred when a survey respondent abandoned the survey. In addition, survey phone calls were recorded with permission of families and one survey per investigator was audited daily by survey managers for survey response accuracy and quality. Families were asked to provide 2 phone numbers and preferred time of calling. Phone calls were made for 5-7 consecutive days, before classifying them as not contactable. The primary outcome measures are described in Table 1. Confounders measured were age of the mother, sex of the baby, whether the baby was premature or not, education of the mother, birth weight less than 2500 gms, parity, delivery type and whether the baby was sent to SNCU.

**Table 1:**
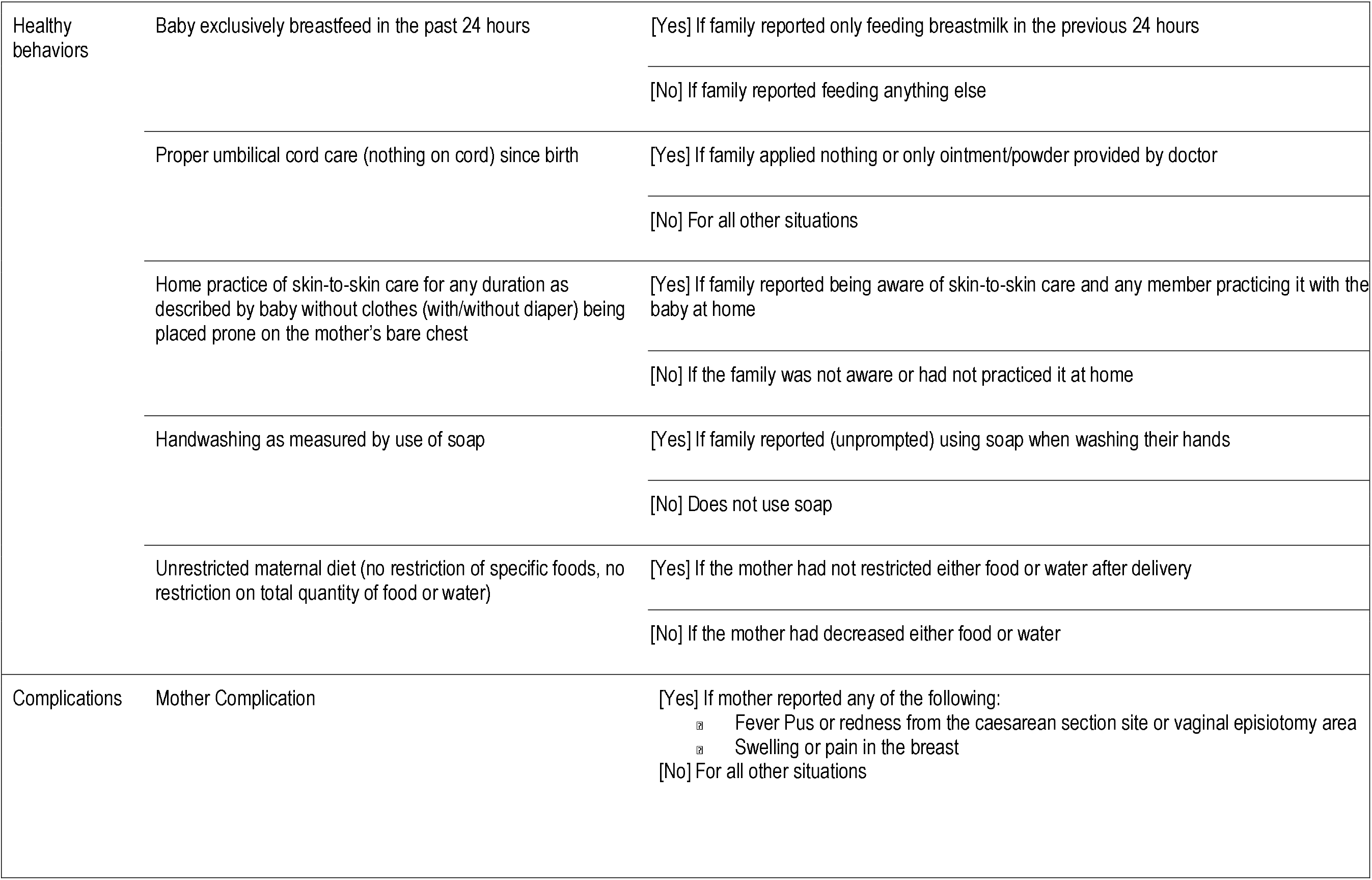
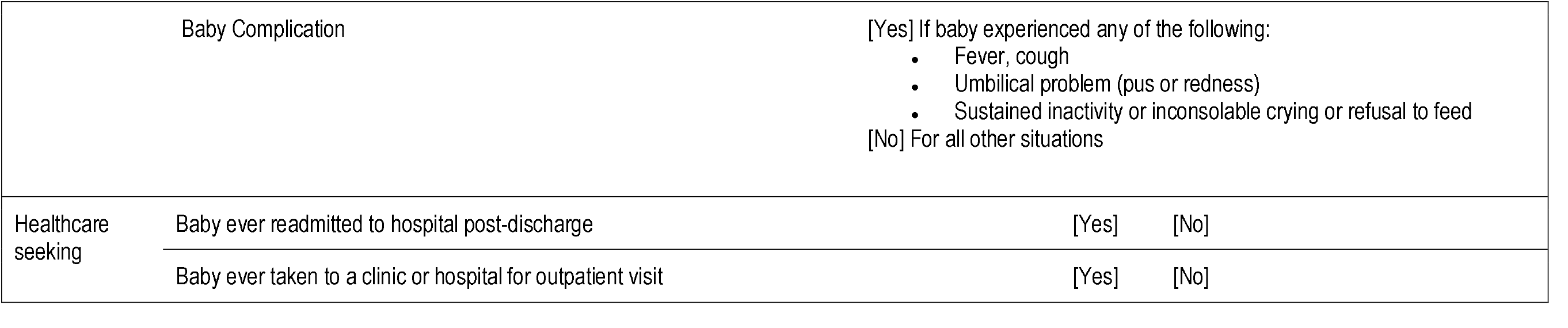
Self-reported outcomes assessed at two-weeks post-delivery

|  |  |  |
| --- | --- | --- |
| Healthy behaviors | Baby exclusively breastfeed in the past 24 hours | [Yes] If family reported only feeding breastmilk in the previous 24 hours |
|  |  | [No] If family reported feeding anything else |
|  | Proper umbilical cord care (nothing on cord) since birth | [Yes] If family applied nothing or only ointment/powder provided by doctor |
|  |  | [No] For all other situations |
|  | Home practice of skin-to-skin care for any duration as described by baby without clothes (with/without diaper) being placed prone on the mother's bare chest | [Yes] If family reported being aware of skin-to-skin care and any member practicing it with the baby at home |
|  |  | [No] If the family was not aware or had not practiced it at home |
|  | Handwashing as measured by use of soap | [Yes] If family reported (unprompted) using soap when washing their hands |
|  |  | [No] Does not use soap |
|  | Unrestricted maternal diet (no restriction of specific foods, no restriction on total quantity of food or water) | [Yes] If the mother had not restricted either food or water after delivery |
|  |  | [No] If the mother had decreased either food or water |
| Complications | Mother Complication | [Yes] If mother reported any of the following:<br>▣ Fever Pus or redness from the caesarean section site or vaginal episiotomy area<br>▣ Swelling or pain in the breast<br>[No] For all other situations |

|  |  |  |  |
| --- | --- | --- | --- |
|  | Baby Complication | [Yes] If baby experienced any of the following:<br>• Fever, cough<br>• Umbilical problem (pus or redness)<br>• Sustained inactivity or inconsolable crying or refusal to feed<br>[No] For all other situations |  |
| Healthcare seeking | Baby ever readmitted to hospital post-discharge | [Yes] | [No] |
|  | Baby ever taken to a clinic or hospital for outpatient visit | [Yes] | [No] |

### Data Analysis

Description of the maternal and child socio-demographic characteristics and confounding variables such as birth weight, gender of baby, whether the delivery was premature, whether it was first pregnancy and whether delivery was through C-section was done using frequencies and percentages. Chi-square test for independence of characteristics between the two intervention phases was used to test significance at 5% level. Logistic regression models with bootstrap standard error were used to assess the effect of intervention on odds of outcome. In the first step, a null model was fit with no predictors. Then models were fit separately for each outcome specified in Table 1 with the intervention group as the primary independent variable and adjusted for confounders. All models were executed for the full sample. Model information efficiency was compared using Akaike’s information criterion (AIC). Effect estimates were in terms of risk ratios which were calculated from odds ratios and significance was reported at 95% confidence.

## RESULTS

We collected 1,507 survey responses from the control families receiving SoC before the launch of the CCP and 3,634 responses from the intervention families in the CCP group. Out of the two selected states, more respondents from Karnataka were in the SoC group (57.4%) as well as the CCP group (68.9%).

Participant demographics are detailed in Table 2. Overall, mother’s age distribution, first pregnancy, type of delivery, baby’s gender, birth weight (<2500g), and preterm status were similar between the two groups. Further, mother’s educational attainment was significantly different (p=0.01) in both phases. However, when combining the education categories, the counts were not statistically different with approximately 1273 (86.3%) of participants in the pre-intervention group with no schooling and 3125 (89%) participants in the post-intervention group with some schooling. In each intervention group, almost fifty percent had attained schooling up to the 6-10th grade category.

**Table 2:**
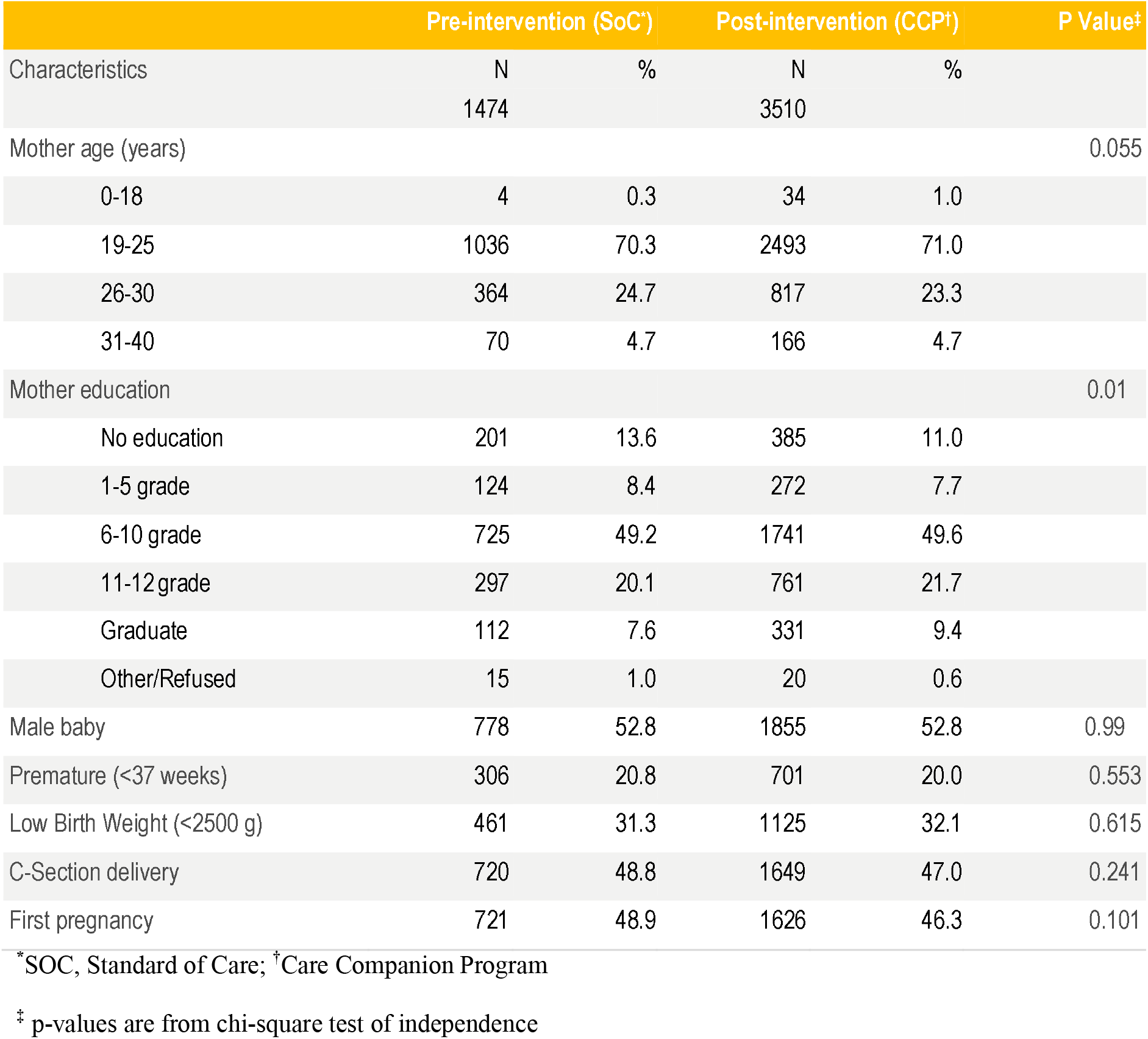
Basic demographic characteristics of participants

The unadjusted and adjusted risk ratios for outcomes in the CCP group compared with the SoC group is shown in Table 3 and the adjusted risk ratios in Figure 1.

**Table 3:** Risk ratios for the post-intervention CCP group as compared to the pre-intervention SoC group

| Outcomes | SoC* N (%) | CCP† N (%) | Risk Ratio |  | 95% CI<sup>§</sup> |
| --- | --- | --- | --- | --- | --- |
|  | 1474 | 3510 | Unadjusted | Adjusted‡ |  |
| Health systems engagement -instructions |  |  |  |  |  |
| Baby care | 482<br>(32.70) | 1268<br>(36.13) | 1.10 | 1.10 | (1.01,1.19) ¶ |
| Warning Signs | 428<br>(29.04) | 1171<br>(33.36) | 1.15 | 1.08 | (0.97,1.18) |
| Healthy behaviors |  |  |  |  |  |
| Exclusive breast feeding | 1232<br>(83.58) | 3076<br>(87.64) | 1.05 | 1.02 | (0.99,1.04) |
| Skin to skin care | 71<br>(4.82) | 324<br>(9.23) | 1.92 | 1.78 | (1.37,2.27) ¶ |
| Dry cord care | 1292<br>(87.65) | 3201<br>(91.20) | 1.04 | 1.04 | (1.02,1.06) ¶ |
| Mother's diet | 449<br>(30.46) | 1083<br>(30.85) | 1.01 | 1.03 | (0.94,1.13) |
| Hand hygiene | 1150<br>(78.02) | 2666<br>(75.95) | 0.97 | 0.98 | (0.94,1.01) |
| Healthcare seeking |  |  |  |  |  |
| Readmission | 70<br>(4.75) | 78<br>(2.22) | 0.47 | 0.44 | (0.31,0.61) ¶ |
| Outpatient visits | 218<br>(14.81) | 658<br>(18.76) | 1.27 | 1.27 | (1.10,1.46) ¶ |

| Outcomes | SoC* N (%) | CCP† N (%) | Risk Ratio |  | 95% CI‡ |
| --- | --- | --- | --- | --- | --- |
| Complications |  |  |  |  |  |
| Newborn | 553<br>(37.52) | 1065<br>(30.34) | 0.81 | 0.84 | (0.76,0.91)¶ |
| Mother | 464<br>(31.48) | 935<br>(26.64) | 0.85 | 0.88 | (0.79,0.97)¶ |

\*SOC, Standard of Care; †CCP, C2are Companion Program
‡ CI, Confidence Interval of adjusted risk ratio.
†Adjusted for age and educational status of mother, gender of baby, prematurity, low birth weight (<2500 grams), gravida and type of delivery
†Significant with confidence interval not including null value

**Figure 1:**
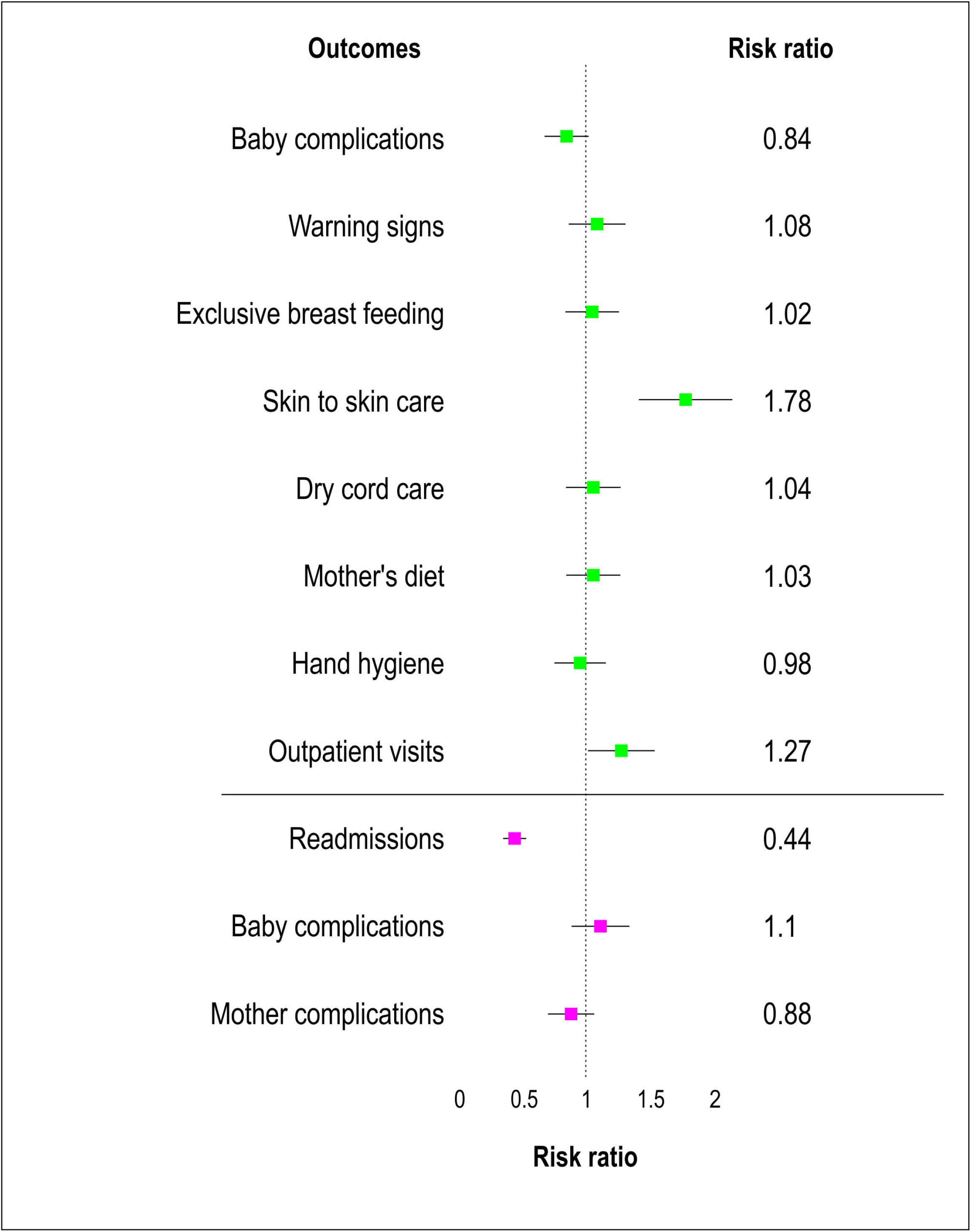

### Health systems engagement-instructions

Ten percent of participants in the CCP group were significantly more likely to have received baby care instructions (RR=1.10 (95 % CI = (1.01, 1.19)). Respondents did not report any differences between the two groups on instructions on baby warning signs.

### Healthy behaviors

Further, we examined healthy behaviors and found, participants in the CCP group were more likely to exclusively breastfeed (2%), 78% more likely to adopt kangaroo care practices, 4% more likely to practice dry cord care, and 3% more likely to have unrestricted diet than those in the SoC group. However, the uptake of hand hygiene was 3% lower in the CCP group. Of these health behaviors, only skin to skin care (adjRR=1.78, 95%CI=1.37,2.27) and dry cord care (adjRR=1.04, 95%CI=1.02,1.06) showed statistically significant improvements (Table 3).

### Healthcare seeking

Health seeking behaviors were significantly improved in the CCP group compared to the SoC group. Outpatient visits were 27% (95% CI=(1.10,1.46)) more likely in the CCP group. Readmissions in the newborn, adjusted for confounders, were 56% (95% CI=(0.31,0.61)) lower in the CCP group.

### Complications

The overall risk of mother or newborn complications reduced significantly in CCP group as compared to SoC group. Risk of newborn complications was reduced by 16% (adjRR=0.84, 95% CI=(0.76,0.91)) and mother complications by 12% (adjRR=0.88, 95% CI=(0.79,0.97)).

## DISCUSSION

### Summary

This study explored the feasibility and effectiveness of a hospital-run, family-focused training program that provides education on multiple evidence-based newborn care practices. Our findings indicate that participation in the in-hospital program was associated with an increase in the uptake of multiple key newborn care practices and health seeking behaviors. The Care Companion Program incorporates postnatal care principles to facilitate the transfer of healthy practices during the critical window of newborn care. Our study showed positive behavior uptake in the right direction for all behaviors except handwashing. However, only behavioral changes associated with skin-to-skin care (RR=1.78, 95%CI=1.37,2.27) and dry cord care (RR=1.04, 95%CI=1.02,1.06) were significant.

### Interpretation

The prevalence of dry cord care in several studies from community settings in India varied from 49% to 72%[4]. Our study showed a much higher prior prevalence at 88%, even before the intervention; this may be because it consisted of only hospital delivered populations. Our skin to skin care was very low in the SoC group at about 5% as compared to other studies in rural India reporting 15%[13]. The reason for this difference is because unlike other studies we measured this practice only after discharge and did not include in-hospital behaviors. We also saw a high uptake of skin-to-skin care as compared to other behaviors. One reason could be that in most hospitals, skin to skin care is promoted only for premature and low birth weight babies, whereas our training included this for all babies. Breastfeeding levels were high at baseline and did not show change with CCP. This may be due to a ceiling effect as most women were already breastfeeding. However, we did not observe this ceiling effect with dry cord care. Importantly, our study showed significant reductions in self-reported neonatal and mother complications with the adjusted risk ratios at 16% and 12%, respectively. Readmissions in the newborn were reduced by 56% after controlling for other variables. Improved and probably timely care seeking, as reported by increased outpatient visits, increased by 27% in the CCP group; this may be associated and partially responsible for the reduced readmissions.

Within India, newborn practices vary and often are related to cultural and religious beliefs. Involvement of entire families allows for easier adoption as additional family members influence uptake[13]. Yet, a comprehensive review of 77 studies in LMICs, of educational strategies for postnatal care, reveals no interventions have considered the involvement of family members beyond the parents[4].The family centered care (FCC) model is useful in discharge planning by equipping families with essential newborn health skills and improving newborn outcomes[14].

The CCP sessions are held with groups of families and the program emphasizes the ability of family members to learn together and practice multiple high-impact health skills. The CCP taps into the collectivist mindset increasing willingness to learn during life events[15] Hospital staff also are able to address any concerns and answer questions during training sessions, which allows for increased engagement and group discussion of difficult care practices. Within overburdened care settings, educational interventions face implementation challenges due to limited availability of healthcare workers. The streamlined CCP design with rotation of staff conducting sessions coupled with group classes alleviates the burden of individual and repetitive counseling. In hospital postnatal education using existing resources can be cost effective compared to community outreach. The in-hospital setting is often the only form of newborn education families receive and prevents missed opportunities to deliver comprehensive information on newborn care[5]. The multi-topic curriculum can be delivered using multiple modalities to target caregiver activation during hospital stays and can improve neonatal outcomes.

### Strengths and Limitations

The primary strengths of our study are its large sample size, measurement of multiple health behaviors and representation of public health settings spanning eleven hospitals in two states. Prior studies evaluating hospital-based postnatal programs have been evaluated in single sites with small sample sizes or only assessed impact of education on single behaviors like breastfeeding. Additionally, this study took place in district hospitals which account for a large proportion of deliveries occurring in government run facilities.

Despite these strengths, there are important limitations to this study. They include the lack of a pre-validated survey and self-reported data. Regarding self-report, there is a chance that people can over-report positive behaviors, but this bias would be expected in both groups. Additionally, our measurement of behaviors could have been improved through in-person assessments and audits of behaviors to verify self-report behavior. However, this was not feasible due to resource constraints.

While some would argue that a quasi-experimental design could be considered a limitation, we disagree. Our study used an historical control group based on the standard of care and in a close timeframe to the intervention. Further, the control group was drawn from the same setting as the interventional arm. Additionally, at the time of our study, no other education improvement or patient counselling efforts were initiated, nor any new programs launched during the duration of the study in any hospital except for one hospital that successfully gained quality improvement accreditation.

Exclusion of mother-baby dyads of families who did not own a phone, were transferred or left the hospital before a data collector could approach them accounted for less than five percent of families. Therefore, we do not consider this to be a significant group. Telephone access was high so almost no families were excluded for this reason. Another point to consider is that while healthy dyads are more likely to leave the hospital quickly, making our population less healthy, the factors pulling it in the opposite direction were sicker dyads being more likely to be transferred. Therefore, we felt the data were balanced in this way.

Most importantly, we conducted the same procedures for data collection before and after the intervention with the same factors affecting both our groups; hence, our results should reflect the population and not be biased.

## Conclusions

This study demonstrated an in-hospital postnatal education program can effectively cover multiple newborn care practices and improve outcomes through a family-centered approach. This is a key finding that can enlighten further efforts to design and evaluate improved postnatal education programs in healthcare settings, particularly in LIMCs. Positive behavior change varied with different behaviors. There is an opportunity for the ongoing program evaluation to allow replication to promote caregiver uptake of newborn care practices following hospital discharge.

The CCP model lends itself for use in LMIC hospital settings, especially where there are supportive family structures for chronic health conditions or where the family is the most consistent factor in the patient’s life and well-being. The simple yet engaging training infrastructure can be integrated into formalized hospital processes as a way to relieve overburdened healthcare systems from downstream complications. The program equips family members with skills that will benefit the patient and enable caregivers to view their roles as an essential component of care delivery.

## Data Availability

The data that support the findings of this study are available from the corresponding author, upon reasonable request.

## Acknowledgements

We would like to thank colleagues from Ariadne Labs, Megan Marx Delaney, Lauren Bobanski, and Katherine Semrau, and Noora colleagues Shirley Yan and Arjun Rangarajan for their review and technical editing. Additionally, we would like to thank Tanmay Singh Pathani, Bhanu Pratap Yadav, Pradeep Kumar K, Anand Kumar, trainers from NHM and data collectors for coordinating data collection and permissions. Revised Standards for Quality Improvement Reporting Excellence (SQUIRE 2.0) publication guidelines guided the manuscript submission

